# Unmasking the determinants of knowledge, attitude and practice of Personal Protective Equipment use among Nepalese farmers using pesticides

**DOI:** 10.1101/2024.05.15.24307406

**Authors:** Reecha Piya, Krishna G.C, Ajay K. Rajbhandari, Sampurna Kakchapati

## Abstract

**Background:** Farmer using commercially available chemical pesticides are at high risk to develop various pesticide-related-illnesses. To mitigate the adverse effect of pesticides, it is crucial for famers to wear Personal Protective Equipment (PPE). This study aimed to assess the factors influencing knowledge, attitude, and practice (KAP) of PPE use among farmers using pesticides in mid-hills of Nepal.

**Methods:** We conducted a cross-sectional study among 256 farmers in Kirtipur Municipality. We collected data using structured questionnaire via telephone. Bi-variate and multivariate logistic regression was done to identify the association between individual, household and farming related characteristic with knowledge, attitude, and practice of PPE use.

**Results:** The study found that the knowledge, attitude, and practice of PPE use was 53.75%, 55.11%, and 47.16%, respectively. The farmers with primary level schooling (AOR= 2.63; 95% CI 1.00-6.88) and secondary level schooling or higher (AOR= 4.19; 95% CI 1.64-0.68) had higher odds of having knowledge on PPE use rather than the farmers with no formal schooling. Farmers of upper lower socioeconomic status had higher odds (AOR=6.40; 95% CI 1.68-24.45) of having positive attitude towards PPE use compared to farmers of upper middle and upper socio-economic status. On the contrary, farmers with good knowledge (AOR=0.20; CI 0.09-0.48) and experience of pesticide-related-illness (AOR=0.29; 95% CI 0.13-0.65) had lower odds of having positive attitude towards PPE use. Married farmers (AOR=10.63; 95% CI 1.11-101.95), farmers using pesticides for more than 10 years (AOR=5.22; 95% CI 1.27-21.43), and farmers using pesticides for 10 hours or more in their lifetime (AOR=4.86; 95% CI 1.63-14.50) had higher odds of wearing basic set of PPE while handling pesticides. However, farmers who were involved in farming for more than 10 years (AOR=0.26; 95% CI 0.07-1.03), farmers who were using pesticide applying methods other than knapsack sprayer (AOR=0.03; 95% CI 0.00-0.34), and farmers having positive attitude towards PPE use (AOR=0.28; 95% CI 0.11-0.70) had lower odds of wearing basic set of PPE while handling pesticides.

**Conclusion:** Farmers in Nepal have a positive outlook on PPE use but multitude of factors including knowledge, farming related factors and socio-economic condition have acted as barrier to its use. Tailored educational programs that not only encompasses knowledge dissemination but also cultivation of positive safety attitudes towards PPE can help increase PPE use.

## Introduction

The advent of pesticides has created a new era in farming, the use of pesticides has helped in creating an increment in crop yields. This increment in crop yields has in turn helped to mitigate global food shortages created due to rapid population growth by controlling the effects of harmful pests in the farms [1]. The use of pesticides is growing and as the trend suggests that in a study conducted between 1945 - 1995 the use of pesticides had doubled every year, especially in the case of developing countries [2]. Similar is the case of Nepal, where the use of pesticides has been increasing year on year since its inception in 1952, a study shows that there is yearly 10-20% growth in use of pesticides in the country to reap short-term and quick benefits from the production of crops [2,3]. It is more prevalent in commercial vegetable farming in Nepal where the use of pesticides is above the national average that is 1.6 kg a.i/ha [4–6].

Pesticides, in general, use a mix of toxic chemicals or biological substances which neutralize pests by killing the population or by repelling them. These toxins not only are harmful to the pests but also to the ecosystem and to human health. The people who are at most risk are the farmers and field workers who are directly exposed to the pesticides during the application process [4]. Many acute and chronic diseases including diarrhoea, nausea, vomiting, asthma, cancer, Parkinson’s diseases, infertility, etc. are directly linked to the exposure to such pesticides[7–9]. Such health conditions are also found to be prevalent among pesticide using farmers in Nepal [10]. It is observed that globally there are more than 3 million (1 million intentional and 2 million unintentional) cases of pesticides poisoning resulting in more than 220,000 deaths every year caused by pesticides [2]. In Nepal also, it was seen that out of 258 cases of acute pesticide poisoning, 6.2% cases were cases of occupational exposure [2].

But these harmful effects caused by pesticides can be mitigated. Studies have shown that the use of Personal Protective Equipment (PPE) can help in reducing health hazards created as a result of direct exposure to pesticides, especially during spraying [11,12]. PPE refers to any items like gloves, safety glasses, bodysuits, or shoes which help to protect various body parts from direct exposure to pesticides [13]. As pesticides can interact with the human body through various sources like skin, eyes or mouth, partial use of PPE cannot be deemed fully protective against pesticides [14]. In Nepal there are instances where farmers were seen using only masks or goggles while dealing with fertilizers, further, there were also instances where farmers were only using long sleeve shirts and long trousers which do not eliminate the harmful effects of pesticides [7,15,16].

Further, there are various social determining factors which influences an individual’s knowledge, attitude, and practice towards PPE use. Factors such as socio-economic status, education, and culture can all have a significant impact [17]. For instance, education and awareness raising activities can increase knowledge and foster positive attitude, eventually influencing the practice of PPE use [18]. In addition, individuals with lower income may have difficulties acquiring PPE, impacting their practice [17]. Likewise, cultural values also play an important role in shaping attitude of PPE and affecting its acceptance and use [19].

Several studies have been conducted in Nepal but most of them have only been concerned with the use of pesticides and its effects on farmers but the conditions relating to use of PPE, however, have been minimal [7,10,15,20–24]. The knowledge, attitude and practices of PPE use and the various factors associated with the use of PPE among farmers and field workers have not been fully explored by studies conducted in the past. Thus, this study primarily focuses on these factors and will help to better understand the risks exposure and the practices relating to use of PPE among farmers while using pesticides.

## Methods

### Study design, population, and setting

We conducted a cross-sectional study among vegetable growing farmers of Kirtipur Municipality of Nepal to identify their knowledge, attitude, practice, and associated factors of PPE use while working with pesticides.

### Sample Size

The sample size for the study was calculated based on a similar study conducted in Nepal. The study reported that the proportion of farmers using combination of protective equipment is 20% [23]. At 95% confidence and 5% margin of error, the minimum sample size required for the study was calculated to be 246. However, to incorporate an anticipated 20% non-response rate, it was inflated to 296.

### Sampling Technique

Considering the situation of COVID-19 pandemic, data collection was done through telephone interviews. The ward-wise list of the farmers along with their contact numbers were collected with the help of local leaders. The list was further refined by selecting only those farmers who were engaged in vegetable farming. From the list thus prepared, 296 farmers were selected randomly using computer-generated random numbers to consider for participation in the study. As data collection was telephone based, at least three attempts were made to contact the selected participants.

### Data collection Tool

The study conducted data collection from July 23, 2021 to September 30, 2021 marking the recruitment period for respondents. A semi-structured questionnaire was developed after extensive literature reviews and consultation with experts. The first section of the questionnaire comprised of demographic characteristics of the participant followed by farming related details. This section was followed by questions related to knowledge, attitude, and practice regarding PPE use. The knowledge section included the harmful effects of pesticides, port of entry and importance of PPE use. The attitude section focused on perception of the farmers regarding perceived susceptibility from pesticides, perceived severity of pesticides, perceived barriers to PPE use and perceived benefits of PPE use. Lastly, the practice section centered around assessing the use of basic set of PPEs in different stages of pesticide handling.

Socio-economic status of the farmers was assessed using Kuppuswamy scale which is based on the scoring of three variables i.e. education of the head of the family of the study participant, occupation of the head of the family of the study participant, and monthly income of the family. Categorization of the socioeconomic status was done based on the total score obtained from the above three variables. Socioeconomic status was categorized as lower (<5), upper-lower (5-10), lower-middle (11-15), upper-middle (16-25), and upper (26-29). The lower and upper-lower category were categorized as poor, lower-middle was categorized as middle, and upper-middle and upper were categorized as rich.

The knowledge section comprised of 19 questions. A score of “1” was attributed for the correct answer and a score of “0” for the incorrect answer. Median score of the questionnaire was calculated and a score below-median score was categorized as poor knowledge while those with equal or above-median score was categorized as good knowledge. Section of attitude included 27 statements with responses documented in the form of a four-point Likert scale: Strongly agree, Agree, Disagree and Strongly Disagree. For identifying the farmers who are practicing the use of PPE, only those farmers who were using mask, gloves, and boot at the same time while mixing the pesticides, spraying the pesticides in the vegetables, cleaning spilled pesticides and disposal of the empty bottles of pesticides were considered as using basic set of PPE use while handling pesticides.

### Operational Definition

Gloves, boots, and mask worn together at a same time was considered as basic set of PPEs. Activities like mixing the pesticides, spraying the pesticides in the vegetables, cleaning spilled pesticides and disposal of the empty bottles of pesticides was considered as pesticide handling.

### Statistical Analysis

A data entry form was prepared in Epi-Info similar to the questionnaire. The response of the participants was extracted from an excel sheet where the data were cleaned and then exported to STATA version 13 for analysis.

Frequency and percentage of the background variables were calculated. For inferential statistics, bivariate analysis of the background variables was carried out with dependent variables. Odds ratio (OR), Confidence Interval (CI), and p-values were obtained to determine important candidate variables for multivariate analysis. The p-values less than 0.05 indicated the significant relationship between variables. The VIF less than 2 with a p-value of less than 0.25 from bivariate analysis were kept in the multivariate analysis. The multivariate analysis of final model was carried out by controlling for both background and mediating variables. From the final model, the p-value of less than 0.05 was considered as a significant association between independent and dependent variable.

### Ethics

Ethical approval was obtained from the Institutional Review Committee (IRC) of Patan Academy of Health Sciences (PAHS)-PAHS IRC Reference PHP2107221558. Before the interview, each participant was briefed about the study and its objectives. Verbal consent was obtained from all the participants. The verbal consent was recorded separately in a mobile phone recorder, and the audio files were copied to a folder in a password protected computer.

## Results

Table 1 shows the socio-demographic and background characteristics of farmers. The median age of the respondent was 40 years. The population of female farmers were nearly twice than male farmers. Most of the farmers were married, comprising 93.68% of the total population. The ethnicity leans heavily towards advantaged ethnic groups (Brahmin, Chettri and Newar) with 78.26% followed by 21.74% from disadvantaged group. More than half of the farmers had completed their secondary schooling or higher (56.52%), followed by those who completed primary school (24.9%), while small proportion (18.58%) had no formal schooling. Regarding the involvement in occupation other than farming, 40.32% of the farmers had other occupation while 59.68% of the farmers were involved exclusively in farming. About the socio-economic status of the farmers, 40.32% were rich, followed by 37.94% middle and 21.74% poor.

**Table 1:** Distribution of respondents by background variables.

| Background Variables | Number (n=253) | Percent (%) |
| --- | --- | --- |
| **Age** |  |  |
| 16-30 | 35 | 13.83 |
| 31-45 | 132 | 52.17 |
| 46-59 | 62 | 24.51 |
| ≥60 | 24 | 9.49 |
| Median age: 40 years |  |  |
| Interquartile Range (IQR): 17 years |  |  |
| **Sex** |  |  |
| Male | 80 | 31.62 |
| Female | 173 | 68.38 |
| **Marital status** |  |  |
| Single | 16 | 6.32 |
| Married | 237 | 93.68 |
| **Relation with Household head** |  |  |
| Household head | 81 | 32.02 |
| Spouse | 158 | 62.45 |
| Others | 14 | 5.53 |
| **Ethnicity** |  |  |
| Advantaged Ethnic Group | 198 | 78.26 |
| Disadvantaged Ethnic Group | 55 | 21.74 |
| **Education of the respondent** |  |  |
| No formal schooling | 47 | 18.58 |
| Primary School | 63 | 24.90 |
| Secondary school and higher | 143 | 56.52 |
| **Involved in Occupation other than farming** |  |  |
| Yes | 102 | 40.32 |
| No | 151 | 59.68 |
| **Socioeconomic Status** |  |  |
| Rich | 102 | 40.32 |
| Middle | 96 | 37.94 |
| Poor | 55 | 21.74 |
| **Households with <5 years children** |  |  |
| Yes | 76 | 30.04 |
| No | 177 | 69.96 |
| **Households with individual aged 60 or older** |  |  |
| Yes | 131 | 51.78 |
| No | 122 | 48.22 |
| **Households with Pregnant women** |  |  |
| Yes | 11 | 4.35 |
| No | 242 | 95.65 |
| **Years involved in farming** |  |  |
| <5 years | 28 | 11.07 |
| 5 to 10 years | 66 | 26.09 |
| >10 years | 159 | 62.85 |
| **Property Relationship** |  |  |
| Property holder | 155 | 61.26 |
| Lease | 98 | 38.74 |
| **Surface area of farmland** |  |  |
| <10952 sq. ft. | 100 | 39.53 |
| ≥10952 sq. ft. | 153 | 60.47 |
| **Purpose of vegetable farming** |  |  |
| Self-consumption | 139 | 54.94 |
| Sell | 114 | 45.06 |
| **Pesticide use** |  |  |
| Yes | 176 | 69.57 |
| No | 77 | 30.43 |
| **Years of pesticides used (N=176)** |  |  |
| <5 years | 78 | 44.32 |
| 5 to 10 years | 78 | 44.32 |
| >10 years | 20 | 11.36 |
| **Vegetable growth in a year (N=176)** |  |  |
| 1 time | 7 | 3.98 |
| 2 times | 99 | 56.25 |
| 3 times | 70 | 39.77 |
| **Number of hours of pesticide exposure in lifetime(N=176)** |  |  |
| <10 hours | 139 | 78.53 |
| ≥10 hours | 38 | 21.47 |
| **Methods of applying pesticides (N=176)** |  |  |
| Knapsack sprayer | 159 | 90.34 |
| Others | 17 | 9.66 |
| **Knowledge (N=253)** |  |  |
| Good | 136 | 53.75 |
| Poor | 117 | 46.25 |
| **Attitude(N=176)** |  |  |
| Positive | 97 | 55.11 |
| Negative | 79 | 44.89 |
| **Training (N=253)** |  |  |
| Yes | 145 | 57.31 |
| No | 108 | 42.69 |
| **Experience of illnesses (N=176)** |  |  |
| Yes | 74 | 42.05 |
| No | 102 | 57.95 |
| **Use of at least one PPE while handling pesticides** |  |  |
| Yes | 167 | 94.89 |
| No | 9 | 5.11 |
| **Basic Set of PPE use while handling pesticides** |  |  |
| Yes | 83 | 47.16 |
| No | 93 | 52.84 |

The majority of farmers were involved in farming for more than 10 years (62.58%) and farming in their own land (61.26%). 54.94% were cultivating vegetables for self-consumption while 45.06% of the farmers were cultivating for commercial purposes. The majority, i.e., 69.57% used pesticides in their vegetables. Among the pesticide users, 44.32% of the respondents were using pesticides for less than 5 years while 44.32% of the respondents were using it for 5 to 10 years, and 11.36% of the respondents were using pesticides for more than 10 years. Regarding the exposure of pesticide in a lifetime, 21.47% of the pesticide users were exposed to pesticides for 10 hours or more in their lifetime. The majority of the farmers (90.34%) used knapsack sprayer for application of pesticides. 53.75% had good knowledge about pesticides and PPE use while 55.11% had positive attitude towards PPE use. 57.31% of the farmers attended some type of training related to agriculture. 42.05% of the pesticide users reported that they had experienced some symptoms related to adverse effects of pesticide use. Among the pesticide users, 94.89% were using at least one form of PPE while handling pesticides while only 47.16% of the farmers were using basic set of PPE while handling pesticides.

Table 2 shows the factors associated with knowledge on PPE use among farmers in both bivariate and multivariate analysis. Education was found to be associated with knowledge of PPE use. The farmers who had completed their primary schooling (AOR= 2.63; 95% CI 1.00-6.88) and secondary level of education or higher (AOR= 4.19; 95% CI 1.64-0.68) had higher odds of having good knowledge about PPE use compared to the farmers who did not have formal education.

**Table 2:**
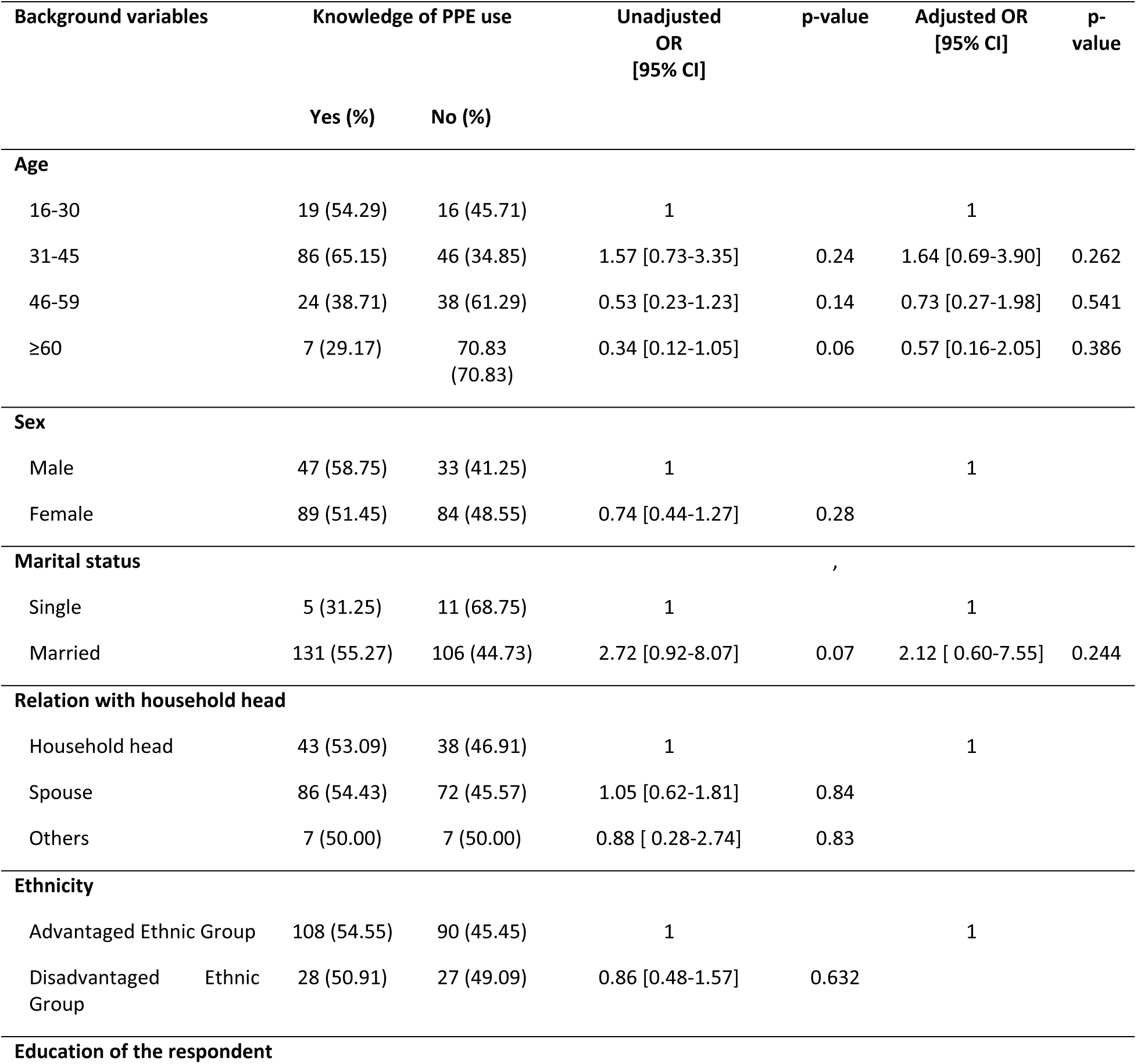
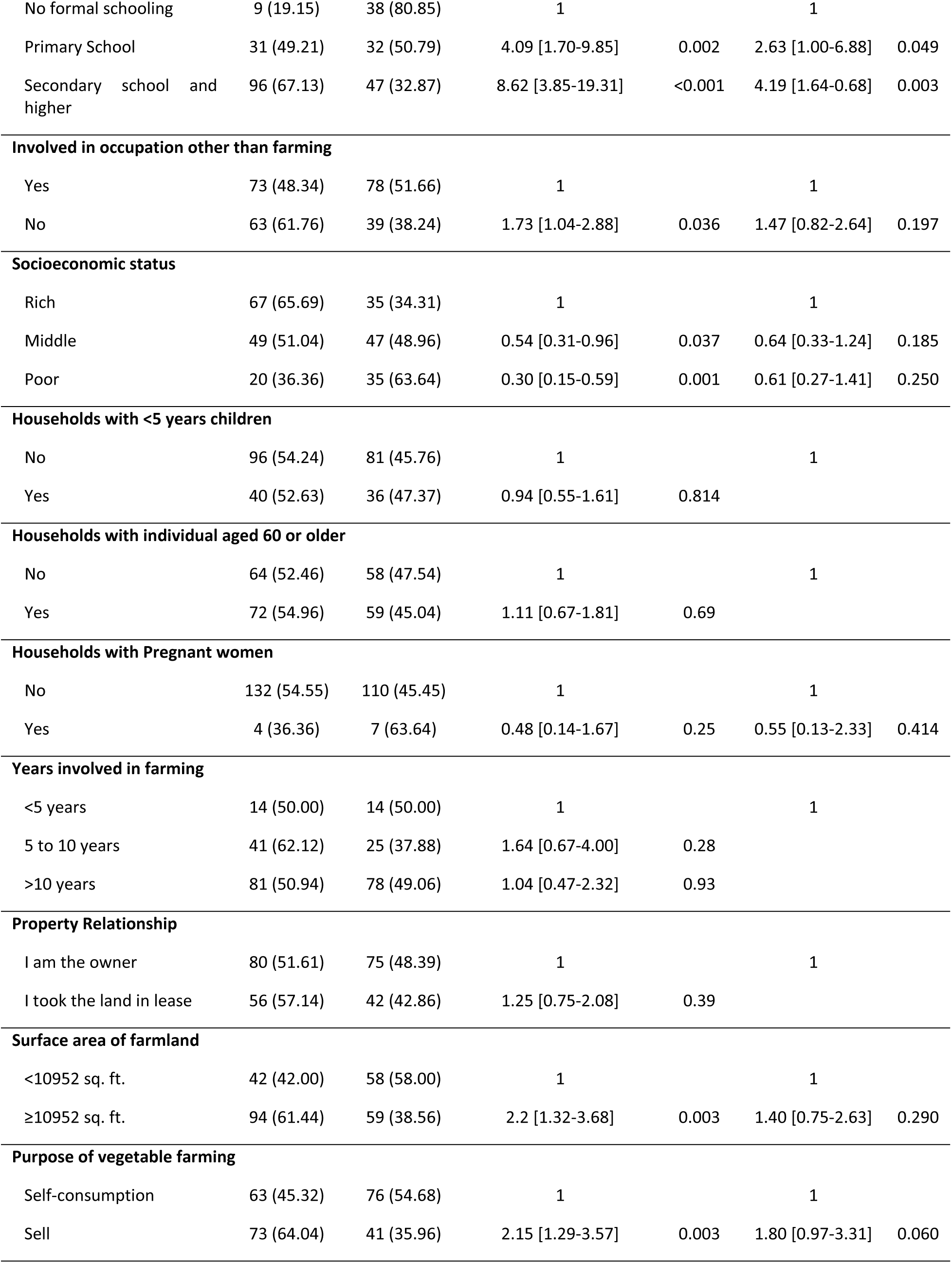
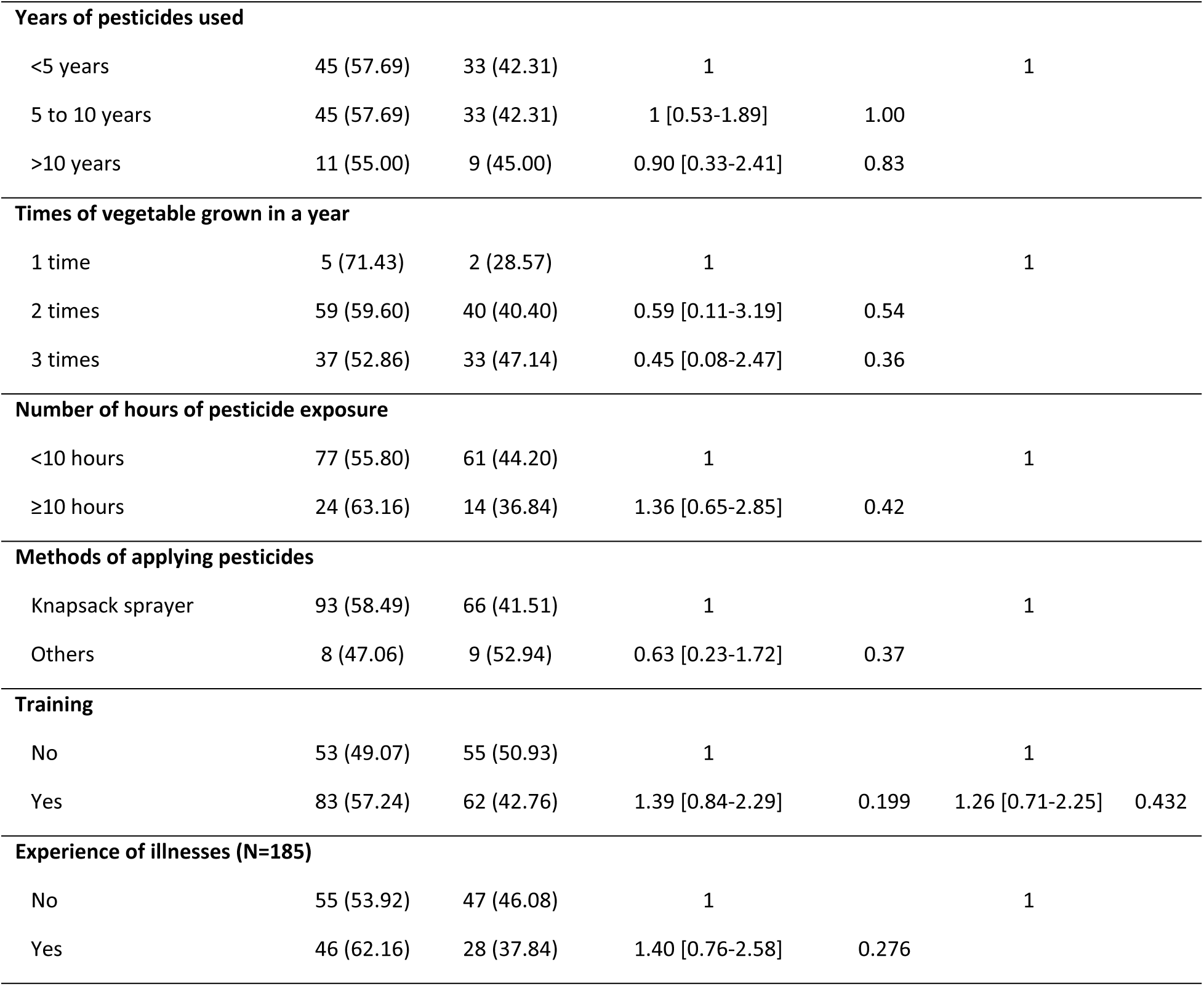
Factors associated with knowledge of PPE use among farmers.

| Background variables |  | Knowledge of PPE use |  | Unadjusted OR<br>[95% CI] | p-value | Adjusted OR<br>[95% CI] | p-value |
| --- | --- | --- | --- | --- | --- | --- | --- |
|  |  | Yes (%) | No (%) |  |  |  |  |
| Age |  |  |  |  |  |  |  |
| 16-30 |  | 19 (54.29) | 16 (45.71) | 1 |  | 1 |  |
| 31-45 |  | 86 (65.15) | 46 (34.85) | 1.57 [0.73-3.35] | 0.24 | 1.64 [0.69-3.90] | 0.262 |
| 46-59 |  | 24 (38.71) | 38 (61.29) | 0.53 [0.23-1.23] | 0.14 | 0.73 [0.27-1.98] | 0.541 |
| ≥60 |  | 7 (29.17) | 70.83 (70.83) | 0.34 [0.12-1.05] | 0.06 | 0.57 [0.16-2.05] | 0.386 |
| Sex |  |  |  |  |  |  |  |
| Male |  | 47 (58.75) | 33 (41.25) | 1 |  | 1 |  |
| Female |  | 89 (51.45) | 84 (48.55) | 0.74 [0.44-1.27] | 0.28 |  |  |
| Marital status |  |  |  |  |  |  |  |
| Single |  | 5 (31.25) | 11 (68.75) | 1 |  | 1 |  |
| Married |  | 131 (55.27) | 106 (44.73) | 2.72 [0.92-8.07] | 0.07 | 2.12 [ 0.60-7.55] | 0.244 |
| Relation with household head |  |  |  |  |  |  |  |
| Household head |  | 43 (53.09) | 38 (46.91) | 1 |  | 1 |  |
| Spouse |  | 86 (54.43) | 72 (45.57) | 1.05 [0.62-1.81] | 0.84 |  |  |
| Others |  | 7 (50.00) | 7 (50.00) | 0.88 [ 0.28-2.74] | 0.83 |  |  |
| Ethnicity |  |  |  |  |  |  |  |
| Advantaged Ethnic Group |  | 108 (54.55) | 90 (45.45) | 1 |  | 1 |  |
| Disadvantaged Ethnic Group |  | 28 (50.91) | 27 (49.09) | 0.86 [0.48-1.57] | 0.632 |  |  |
| Education of the respondent |  |  |  |  |  |  |  |

|  |  |  |  |  |  |  |
| --- | --- | --- | --- | --- | --- | --- |
| No formal schooling | 9 (19.15) | 38 (80.85) | 1 |  | 1 |  |
| Primary School | 31 (49.21) | 32 (50.79) | 4.09 [1.70-9.85] | 0.002 | 2.63 [1.00-6.88] | 0.049 |
| Secondary school and higher | 96 (67.13) | 47 (32.87) | 8.62 [3.85-19.31] | <0.001 | 4.19 [1.64-0.68] | 0.003 |
| **Involved in occupation other than farming** |  |  |  |  |  |  |
| Yes | 73 (48.34) | 78 (51.66) | 1 |  | 1 |  |
| No | 63 (61.76) | 39 (38.24) | 1.73 [1.04-2.88] | 0.036 | 1.47 [0.82-2.64] | 0.197 |
| **Socioeconomic status** |  |  |  |  |  |  |
| Rich | 67 (65.69) | 35 (34.31) | 1 |  | 1 |  |
| Middle | 49 (51.04) | 47 (48.96) | 0.54 [0.31-0.96] | 0.037 | 0.64 [0.33-1.24] | 0.185 |
| Poor | 20 (36.36) | 35 (63.64) | 0.30 [0.15-0.59] | 0.001 | 0.61 [0.27-1.41] | 0.250 |
| **Households with <5 years children** |  |  |  |  |  |  |
| No | 96 (54.24) | 81 (45.76) | 1 |  | 1 |  |
| Yes | 40 (52.63) | 36 (47.37) | 0.94 [0.55-1.61] | 0.814 |  |  |
| **Households with individual aged 60 or older** |  |  |  |  |  |  |
| No | 64 (52.46) | 58 (47.54) | 1 |  | 1 |  |
| Yes | 72 (54.96) | 59 (45.04) | 1.11 [0.67-1.81] | 0.69 |  |  |
| **Households with Pregnant women** |  |  |  |  |  |  |
| No | 132 (54.55) | 110 (45.45) | 1 |  | 1 |  |
| Yes | 4 (36.36) | 7 (63.64) | 0.48 [0.14-1.67] | 0.25 | 0.55 [0.13-2.33] | 0.414 |
| **Years involved in farming** |  |  |  |  |  |  |
| <5 years | 14 (50.00) | 14 (50.00) | 1 |  | 1 |  |
| 5 to 10 years | 41 (62.12) | 25 (37.88) | 1.64 [0.67-4.00] | 0.28 |  |  |
| >10 years | 81 (50.94) | 78 (49.06) | 1.04 [0.47-2.32] | 0.93 |  |  |
| **Property Relationship** |  |  |  |  |  |  |
| I am the owner | 80 (51.61) | 75 (48.39) | 1 |  | 1 |  |
| I took the land in lease | 56 (57.14) | 42 (42.86) | 1.25 [0.75-2.08] | 0.39 |  |  |
| **Surface area of farmland** |  |  |  |  |  |  |
| <10952 sq. ft. | 42 (42.00) | 58 (58.00) | 1 |  | 1 |  |
| ≥10952 sq. ft. | 94 (61.44) | 59 (38.56) | 2.2 [1.32-3.68] | 0.003 | 1.40 [0.75-2.63] | 0.290 |
| **Purpose of vegetable farming** |  |  |  |  |  |  |
| Self-consumption | 63 (45.32) | 76 (54.68) | 1 |  | 1 |  |
| Sell | 73 (64.04) | 41 (35.96) | 2.15 [1.29-3.57] | 0.003 | 1.80 [0.97-3.31] | 0.060 |
| **Years of pesticides used** |  |  |  |  |  |  |
| <5 years | 45 (57.69) | 33 (42.31) | 1 |  | 1 |  |
| 5 to 10 years | 45 (57.69) | 33 (42.31) | 1 [0.53-1.89] | 1.00 |  |  |
| >10 years | 11 (55.00) | 9 (45.00) | 0.90 [0.33-2.41] | 0.83 |  |  |
| **Times of vegetable grown in a year** |  |  |  |  |  |  |
| 1 time | 5 (71.43) | 2 (28.57) | 1 |  | 1 |  |
| 2 times | 59 (59.60) | 40 (40.40) | 0.59 [0.11-3.19] | 0.54 |  |  |
| 3 times | 37 (52.86) | 33 (47.14) | 0.45 [0.08-2.47] | 0.36 |  |  |
| **Number of hours of pesticide exposure** |  |  |  |  |  |  |
| <10 hours | 77 (55.80) | 61 (44.20) | 1 |  | 1 |  |
| ≥10 hours | 24 (63.16) | 14 (36.84) | 1.36 [0.65-2.85] | 0.42 |  |  |
| **Methods of applying pesticides** |  |  |  |  |  |  |
| Knapsack sprayer | 93 (58.49) | 66 (41.51) | 1 |  | 1 |  |
| Others | 8 (47.06) | 9 (52.94) | 0.63 [0.23-1.72] | 0.37 |  |  |
| **Training** |  |  |  |  |  |  |
| No | 53 (49.07) | 55 (50.93) | 1 |  | 1 |  |
| Yes | 83 (57.24) | 62 (42.76) | 1.39 [0.84-2.29] | 0.199 | 1.26 [0.71-2.25] | 0.432 |
| **Experience of illnesses (N=185)** |  |  |  |  |  |  |
| No | 55 (53.92) | 47 (46.08) | 1 |  | 1 |  |
| Yes | 46 (62.16) | 28 (37.84) | 1.40 [0.76-2.58] | 0.276 |  |  |

Table 3 shows the factors associated with attitude towards PPE use among farmers in both bivariate and multivariate analysis. Socioeconomic status, knowledge with the farmers and experience of illness were found to be associated with attitude of farmers towards PPE use. The farmers belonging to the upper lower (IV) category (AOR=6.40; 95% CI 1.68-24.45) had higher odds of having positive attitude towards PPE use than the farmers belonging to upper and upper middle category. However, the farmers with good knowledge of PPE use had lower odds of having positive attitude towards PPE use (AOR=0.20; CI 0.09-0.48) than the farmers with poor knowledge. Likewise, the farmers who had previous experience of illness related to pesticide use had lower odds of having positive attitude towards PPE use (AOR=0.29; 95% CI 0.13-0.65) than those farmers who did not experience any illnesses.

**Table 3:**
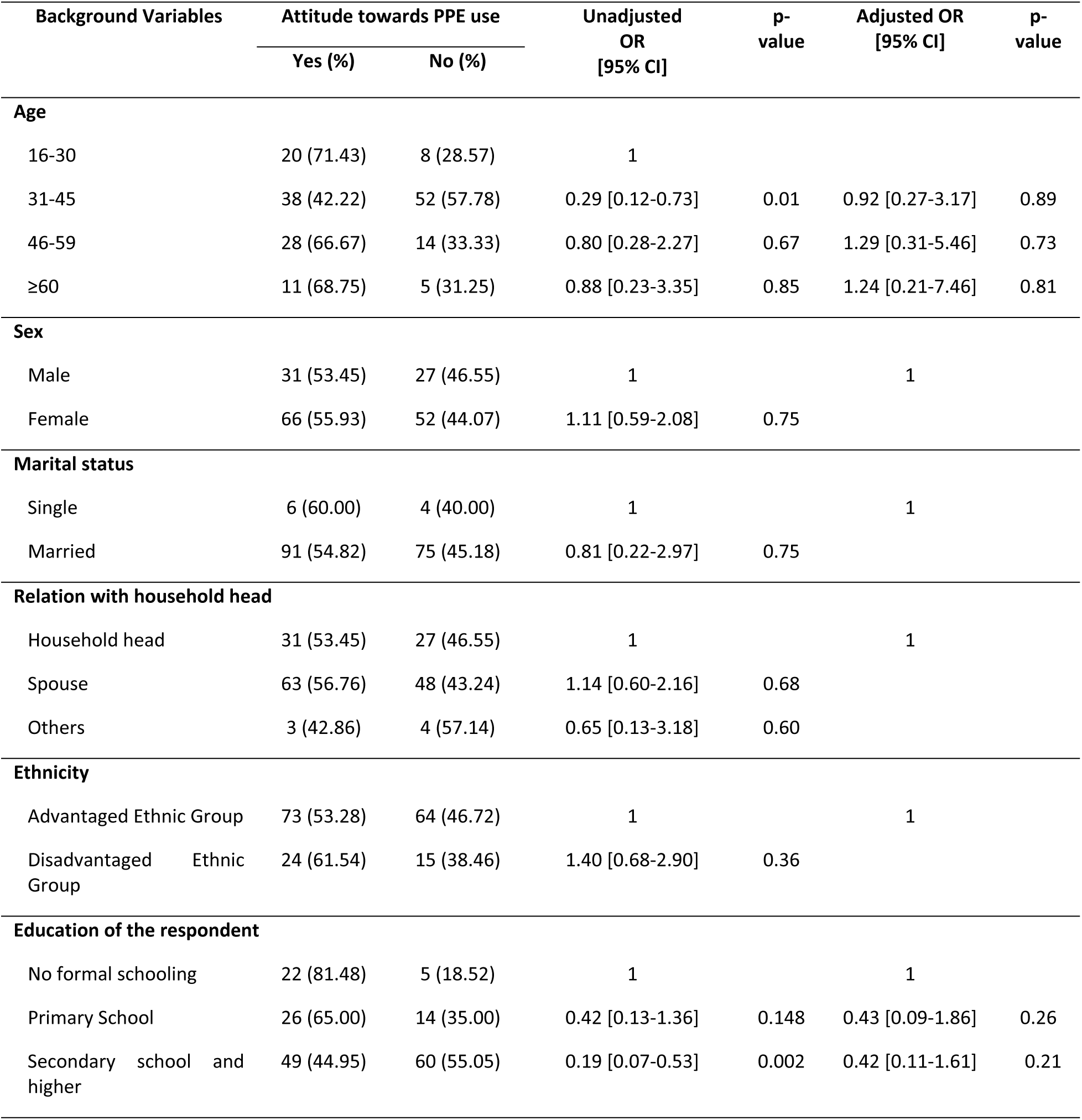
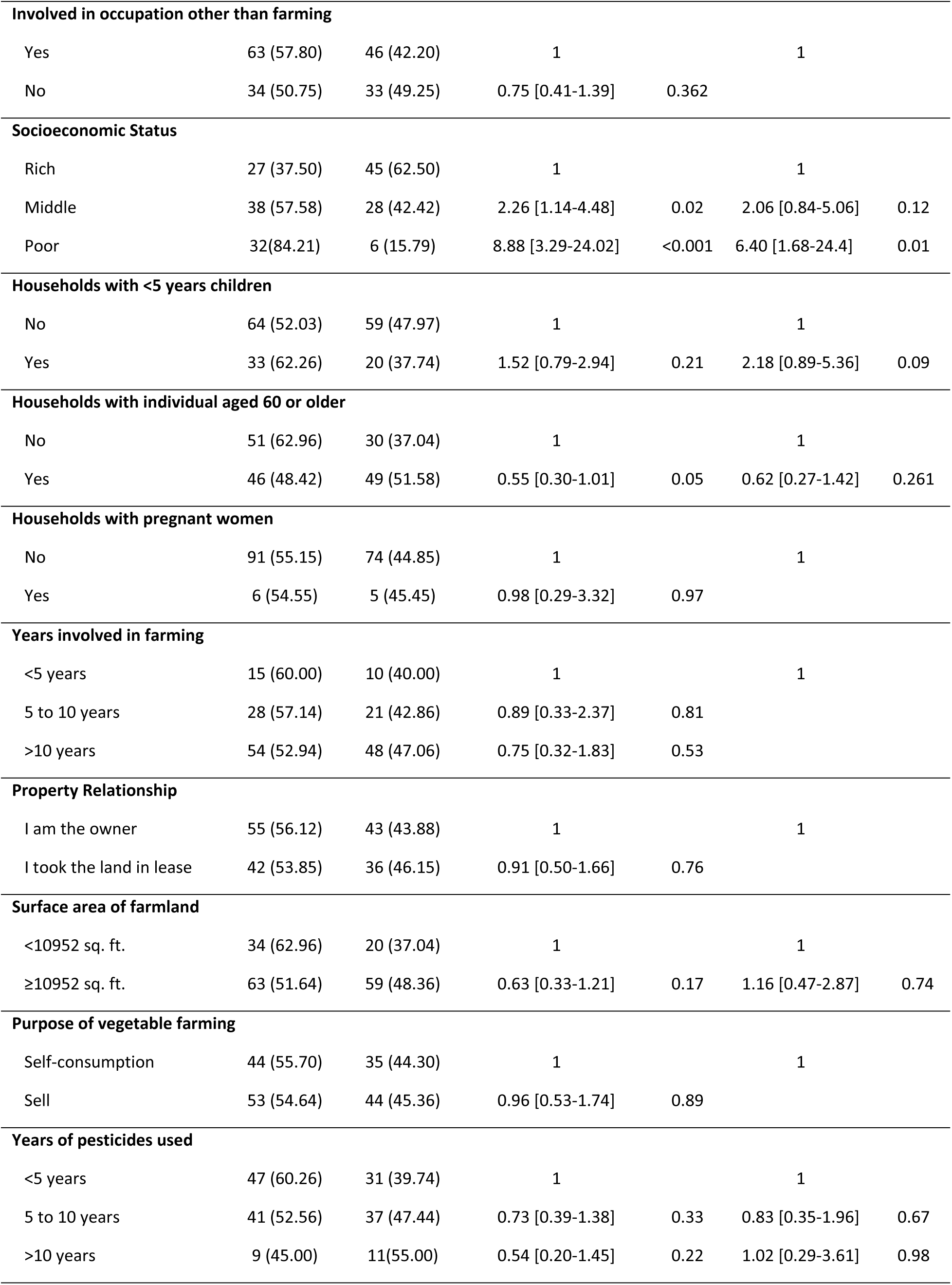
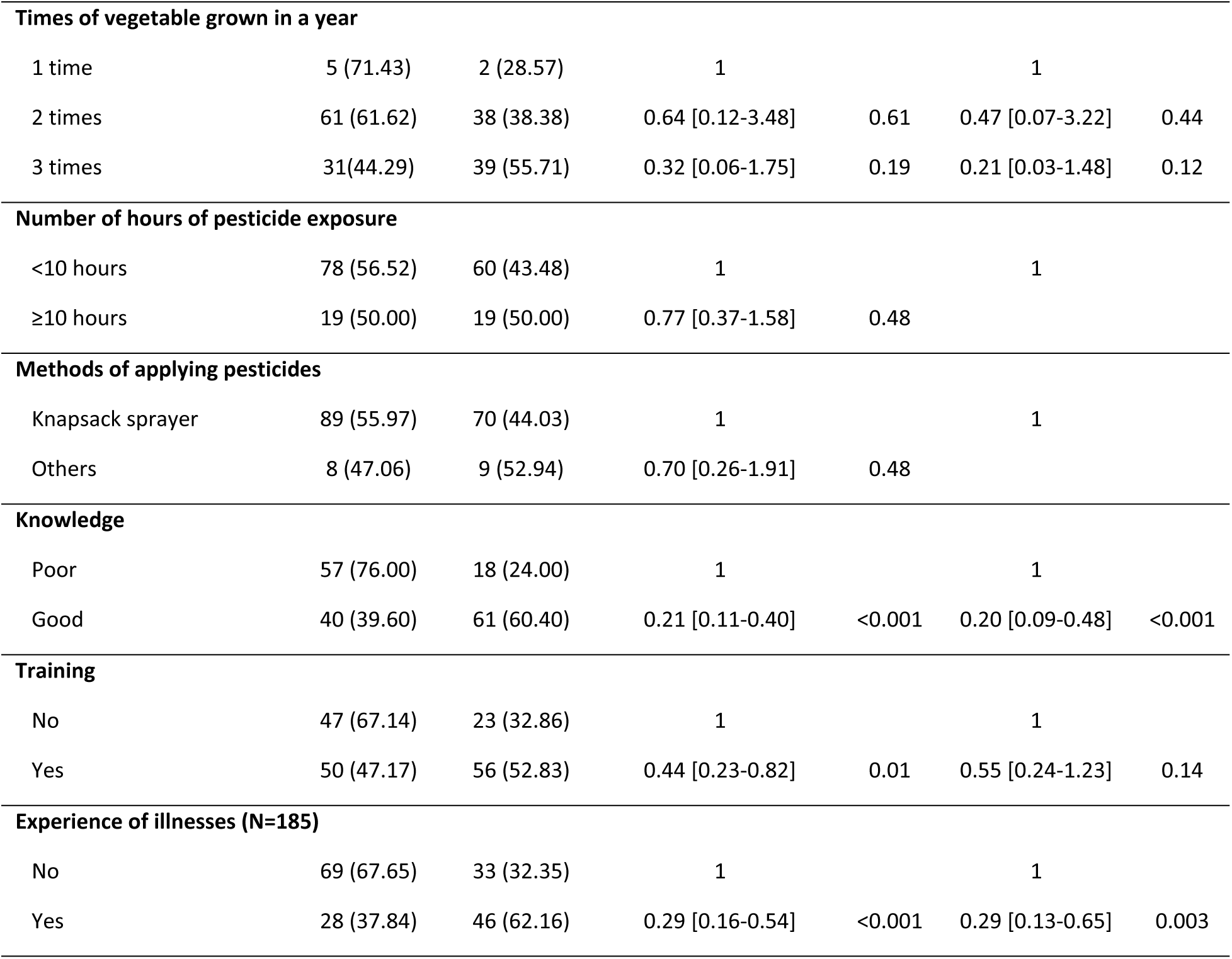
Factors associated with Attitude towards PPE use among farmers.

Table 4 shows the factors associated with practice on PPE use among farmers in both bivariate and multivariate analysis. Marital status, years involved in farming, years of pesticide use, number of hours of pesticide exposure, methods of applying pesticides, and attitude towards PPE use were found to be associated with practice of PPE use.

**Table 4:**
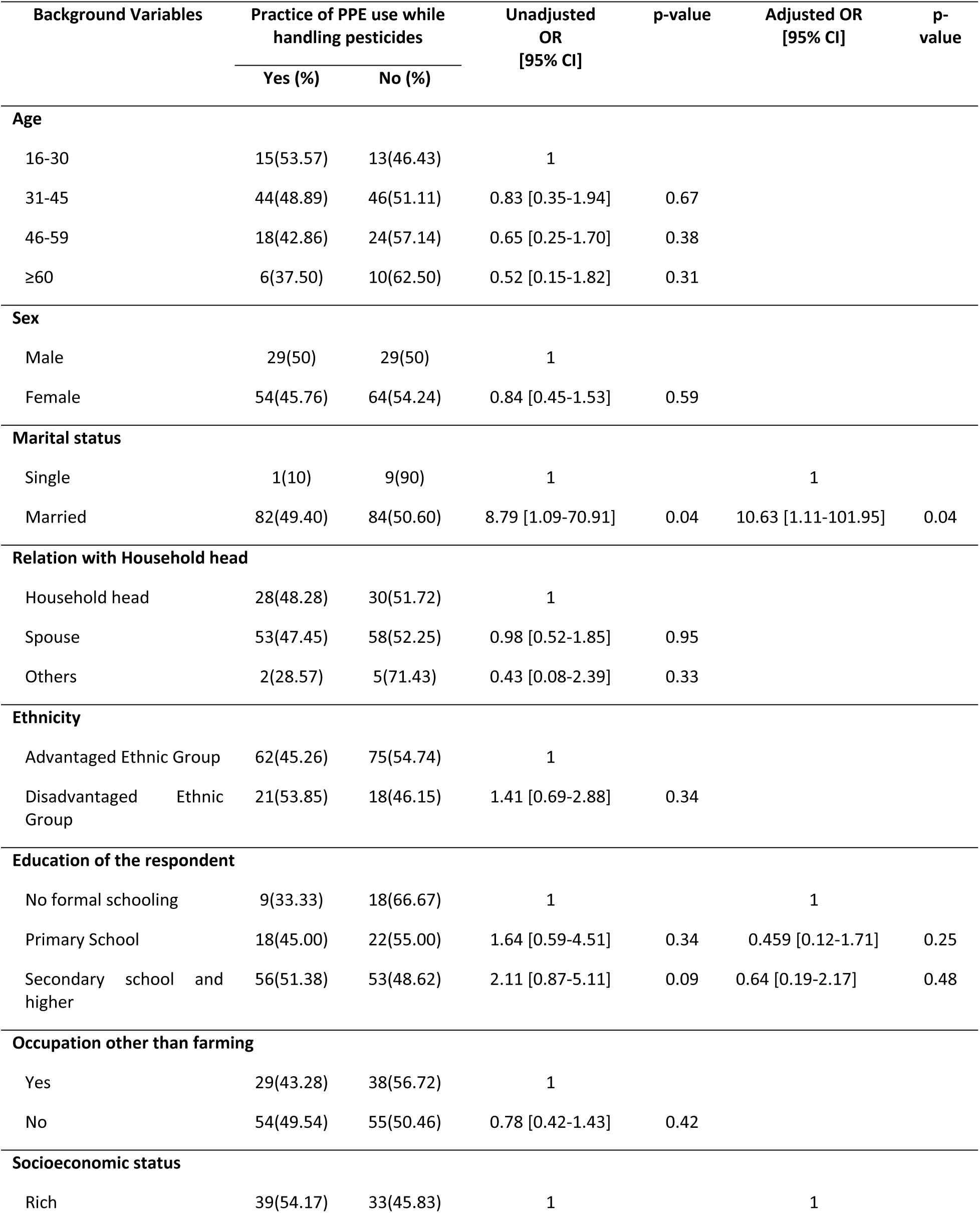
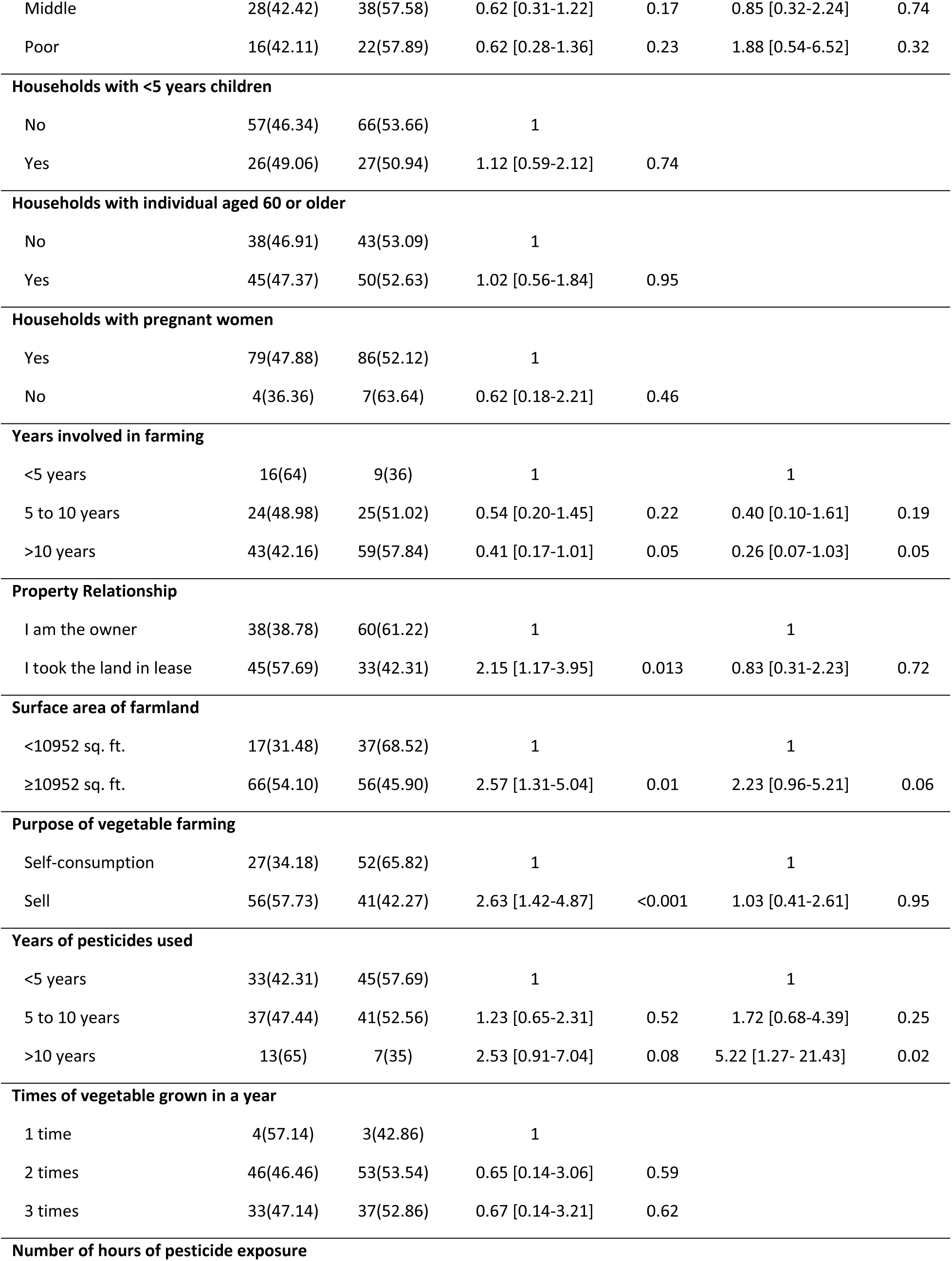
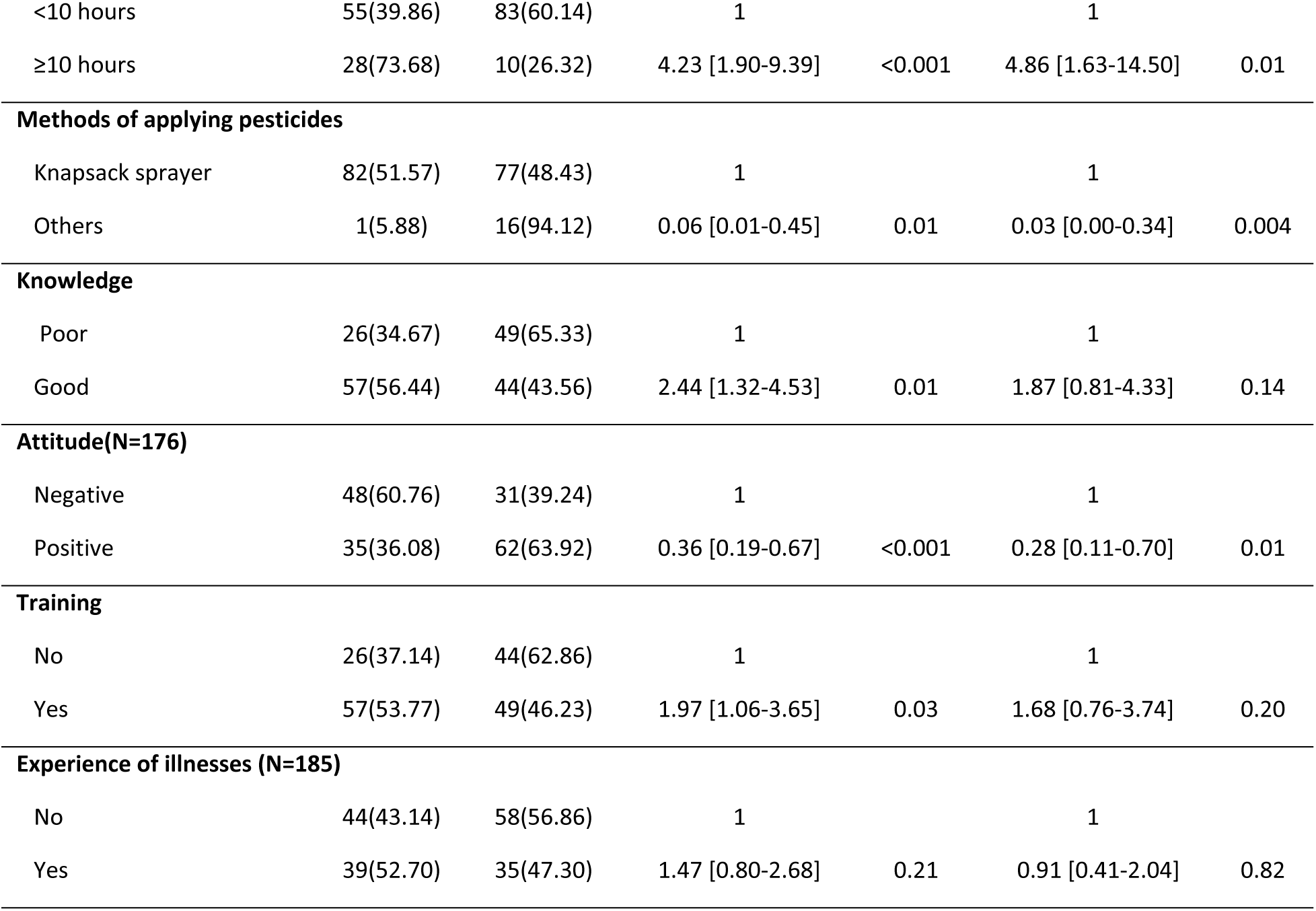
Factors associated with Practice of PPE use among farmers.

| Background Variables |  | Practice of PPE use while handling pesticides |  | Unadjusted OR [95% CI] | p-value | Adjusted OR [95% CI] | p-value |
| --- | --- | --- | --- | --- | --- | --- | --- |
|  |  | Yes (%) | No (%) |  |  |  |  |
| Age |  |  |  |  |  |  |  |
| 16-30 |  | 15(53.57) | 13(46.43) | 1 |  |  |  |
| 31-45 |  | 44(48.89) | 46(51.11) | 0.83 [0.35-1.94] | 0.67 |  |  |
| 46-59 |  | 18(42.86) | 24(57.14) | 0.65 [0.25-1.70] | 0.38 |  |  |
| ≥60 |  | 6(37.50) | 10(62.50) | 0.52 [0.15-1.82] | 0.31 |  |  |
| Sex |  |  |  |  |  |  |  |
| Male |  | 29(50) | 29(50) | 1 |  |  |  |
| Female |  | 54(45.76) | 64(54.24) | 0.84 [0.45-1.53] | 0.59 |  |  |
| Marital status |  |  |  |  |  |  |  |
| Single |  | 1(10) | 9(90) | 1 |  | 1 |  |
| Married |  | 82(49.40) | 84(50.60) | 8.79 [1.09-70.91] | 0.04 | 10.63 [1.11-101.95] | 0.04 |
| Relation with Household head |  |  |  |  |  |  |  |
| Household head |  | 28(48.28) | 30(51.72) | 1 |  |  |  |
| Spouse |  | 53(47.45) | 58(52.25) | 0.98 [0.52-1.85] | 0.95 |  |  |
| Others |  | 2(28.57) | 5(71.43) | 0.43 [0.08-2.39] | 0.33 |  |  |
| Ethnicity |  |  |  |  |  |  |  |
| Advantaged Ethnic Group |  | 62(45.26) | 75(54.74) | 1 |  |  |  |
| Disadvantaged Ethnic Group |  | 21(53.85) | 18(46.15) | 1.41 [0.69-2.88] | 0.34 |  |  |
| Education of the respondent |  |  |  |  |  |  |  |
| No formal schooling |  | 9(33.33) | 18(66.67) | 1 |  | 1 |  |
| Primary School |  | 18(45.00) | 22(55.00) | 1.64 [0.59-4.51] | 0.34 | 0.459 [0.12-1.71] | 0.25 |
| Secondary school and higher |  | 56(51.38) | 53(48.62) | 2.11 [0.87-5.11] | 0.09 | 0.64 [0.19-2.17] | 0.48 |
| Occupation other than farming |  |  |  |  |  |  |  |
| Yes |  | 29(43.28) | 38(56.72) | 1 |  |  |  |
| No |  | 54(49.54) | 55(50.46) | 0.78 [0.42-1.43] | 0.42 |  |  |
| Socioeconomic status |  |  |  |  |  |  |  |
| Rich |  | 39(54.17) | 33(45.83) | 1 |  | 1 |  |

|  |  |  |  |  |  |  |
| --- | --- | --- | --- | --- | --- | --- |
| Middle | 28(42.42) | 38(57.58) | 0.62 [0.31-1.22] | 0.17 | 0.85 [0.32-2.24] | 0.74 |
| Poor | 16(42.11) | 22(57.89) | 0.62 [0.28-1.36] | 0.23 | 1.88 [0.54-6.52] | 0.32 |
| **Households with <5 years children** |  |  |  |  |  |  |
| No | 57(46.34) | 66(53.66) | 1 |  |  |  |
| Yes | 26(49.06) | 27(50.94) | 1.12 [0.59-2.12] | 0.74 |  |  |
| **Households with individual aged 60 or older** |  |  |  |  |  |  |
| No | 38(46.91) | 43(53.09) | 1 |  |  |  |
| Yes | 45(47.37) | 50(52.63) | 1.02 [0.56-1.84] | 0.95 |  |  |
| **Households with pregnant women** |  |  |  |  |  |  |
| Yes | 79(47.88) | 86(52.12) | 1 |  |  |  |
| No | 4(36.36) | 7(63.64) | 0.62 [0.18-2.21] | 0.46 |  |  |
| **Years involved in farming** |  |  |  |  |  |  |
| <5 years | 16(64) | 9(36) | 1 |  | 1 |  |
| 5 to 10 years | 24(48.98) | 25(51.02) | 0.54 [0.20-1.45] | 0.22 | 0.40 [0.10-1.61] | 0.19 |
| >10 years | 43(42.16) | 59(57.84) | 0.41 [0.17-1.01] | 0.05 | 0.26 [0.07-1.03] | 0.05 |
| **Property Relationship** |  |  |  |  |  |  |
| I am the owner | 38(38.78) | 60(61.22) | 1 |  | 1 |  |
| I took the land in lease | 45(57.69) | 33(42.31) | 2.15 [1.17-3.95] | 0.013 | 0.83 [0.31-2.23] | 0.72 |
| **Surface area of farmland** |  |  |  |  |  |  |
| <10952 sq. ft. | 17(31.48) | 37(68.52) | 1 |  | 1 |  |
| ≥10952 sq. ft. | 66(54.10) | 56(45.90) | 2.57 [1.31-5.04] | 0.01 | 2.23 [0.96-5.21] | 0.06 |
| **Purpose of vegetable farming** |  |  |  |  |  |  |
| Self-consumption | 27(34.18) | 52(65.82) | 1 |  | 1 |  |
| Sell | 56(57.73) | 41(42.27) | 2.63 [1.42-4.87] | <0.001 | 1.03 [0.41-2.61] | 0.95 |
| **Years of pesticides used** |  |  |  |  |  |  |
| <5 years | 33(42.31) | 45(57.69) | 1 |  | 1 |  |
| 5 to 10 years | 37(47.44) | 41(52.56) | 1.23 [0.65-2.31] | 0.52 | 1.72 [0.68-4.39] | 0.25 |
| >10 years | 13(65) | 7(35) | 2.53 [0.91-7.04] | 0.08 | 5.22 [1.27- 21.43] | 0.02 |
| **Times of vegetable grown in a year** |  |  |  |  |  |  |
| 1 time | 4(57.14) | 3(42.86) | 1 |  |  |  |
| 2 times | 46(46.46) | 53(53.54) | 0.65 [0.14-3.06] | 0.59 |  |  |
| 3 times | 33(47.14) | 37(52.86) | 0.67 [0.14-3.21] | 0.62 |  |  |
| **Number of hours of pesticide exposure** |  |  |  |  |  |  |
| <10 hours | 55(39.86) | 83(60.14) | 1 |  | 1 |  |
| ≥10 hours | 28(73.68) | 10(26.32) | 4.23 [1.90-9.39] | <0.001 | 4.86 [1.63-14.50] | 0.01 |
| **Methods of applying pesticides** |  |  |  |  |  |  |
| Knapsack sprayer | 82(51.57) | 77(48.43) | 1 |  | 1 |  |
| Others | 1(5.88) | 16(94.12) | 0.06 [0.01-0.45] | 0.01 | 0.03 [0.00-0.34] | 0.004 |
| **Knowledge** |  |  |  |  |  |  |
| Poor | 26(34.67) | 49(65.33) | 1 |  | 1 |  |
| Good | 57(56.44) | 44(43.56) | 2.44 [1.32-4.53] | 0.01 | 1.87 [0.81-4.33] | 0.14 |
| **Attitude(N=176)** |  |  |  |  |  |  |
| Negative | 48(60.76) | 31(39.24) | 1 |  | 1 |  |
| Positive | 35(36.08) | 62(63.92) | 0.36 [0.19-0.67] | <0.001 | 0.28 [0.11-0.70] | 0.01 |
| **Training** |  |  |  |  |  |  |
| No | 26(37.14) | 44(62.86) | 1 |  | 1 |  |
| Yes | 57(53.77) | 49(46.23) | 1.97 [1.06-3.65] | 0.03 | 1.68 [0.76-3.74] | 0.20 |
| **Experience of illnesses (N=185)** |  |  |  |  |  |  |
| No | 44(43.14) | 58(56.86) | 1 |  | 1 |  |
| Yes | 39(52.70) | 35(47.30) | 1.47 [0.80-2.68] | 0.21 | 0.91 [0.41-2.04] | 0.82 |

The farmers who were married had higher odds of wearing PPE while working with pesticides (AOR=10.63; 95% CI 1.11-101.95) compared to unmarried farmers. The farmers who were using pesticides in their vegetables for more than 10 years were had higher odds of wearing PPE while working with pesticides (AOR=5.22; 95% CI 1.27-21.43) than those farmers who were using pesticides for less than 5 years. The farmers who were exposed to pesticides for 10 hours or more in their lifetime had higher odds of wearing PPE while working with pesticides (AOR=4.86; 95% CI 1.63-14.50) than the farmers who were exposed to pesticides for less than 10 hours in their lifetime. In contrast, the farmers who were involved in vegetable farming for more than 10 years had lower odds of using PPE while working with pesticides (AOR=0.26; 95% CI 0.07-1.03) than those farmers who had been involved in farming for less than 5 years. The farmers who were using other pesticide application methods other than knapsack sprayer had lower odds of wearing PPE while working with pesticides (AOR=0.03; 95% CI 0.00-0.34) than those farmers using knapsack sprayer. The farmers who had positive attitude towards PPE use had lower odds of wearing PPE while working with pesticides (AOR=0.28; 95% CI 0.11-0.70) than those having negative attitude towards PPE use.

## Discussion

To the best of our understanding, this is the first study determining the knowledge, attitude, practice, and associated factors of PPE use among farmers in Nepal, as previously most of them had limited themselves to the safe handling of pesticides and illnesses resulting from pesticide use [7,10,20–24]. This study also found that more than half of farmers had adequate knowledge on the importance of PPE use and adverse effects of pesticides. This is quite different from other studies conducted in Nepal which had reported that more than 80% of the farmers possessed knowledge related to adverse health effects of pesticides, while, a study conducted in Thailand had reported a much lower knowledge ratio i.e. 22.8%, the difference in the result might be because most of these studies concentrated themselves with the study of harmful effects of pesticides, while our study also focused on the use of PPE [7,22,25]. Also, while measuring the attitude of farmers towards use of PPE in this study, the findings are quite different from the findings revealed by a study conducted in Iran which reported less than half of the farmer’s intended to use PPE while handling pesticides [26].

Pesticides can enter the body through various routes like ingestion, inhalation, and skin absorption thus wearing a combination of PPE is crucial in order to minimize this exposure [9].Our study showed that, almost all the farmers were wearing at least one form of PPE while handling pesticides but when it came to wearing a combination of PPE, less than half of the population were found to be wearing mask, gloves and boot at the same time which is considered as the basic set of PPE, the proportion was similar to a study conducted in Brazil [27]. This prevalence revealed by our study is higher in Nepal, than in countries like Ethiopia (10%) and Thailand (6%) [25,28]. Further, other studies conducted in other parts of Nepal in various years showed a comparatively lower proportion of PPE use as compared to our study [20,22,23]. The possible reason behind this might be due to the fact that our study was conducted post COVID while other studies were conducted before the pandemic [20,22,23]. The COVID pandemic heightened the awareness about PPE for disease prevention and led to the heightened demand and usage of PPE. The increase in production and distribution led to an increased availability, leading to an increased proportion of PPE use among farmers in our study [29]. However, it is contradicting, as our study also showed that more than half of the population of farmers do not use basic set of PPE while handling pesticides, which implies that farmers in Nepal are still at high risk of developing various pesticide-related-illnesses.

Further, association between education levels and knowledge regarding PPE use among farmers also constitutes a notable finding in this study. This association is in line with previous studies highlighting the influential role of education in shaping safety practices in various occupational settings [30,31]. Higher levels of education are often linked with enhanced access to formalized information channels, including agricultural training programs and workshops. These avenues offer a platform for disseminating critical safety information, including the proper use of PPE. Additionally, farmers with higher educational attainment may possess superior cognitive abilities, potentially affording them a greater capacity to comprehend and retain safety-related information [30,31].

In addition to that, this study also showed a significant association between lower socioeconomic status and attitudes towards PPE use among farmers. This finding is in alignment with existing literature, which underscores the intricate interplay between economic resources and safety practices in occupational settings [32,33]. Farmers with limited financial means may encounter barriers in adopting favorable attitudes towards PPE use [33]. Financial constraints often hinder their capacity to invest in safety equipment, diverting resources towards more immediate needs. Moreover, this association may be exacerbated by a lack of access to formalized safety training programs and information materials, which are crucial for cultivating positive attitudes towards safety practices. Moreover, a noteworthy observation emerged from the study, indicating that farmers who reported instances of illness exhibited lower odds of holding favorable attitudes towards the use of PPE. This finding resonates with prior research emphasizing the multifaceted relationship between health status and safety practices in agricultural contexts [25,32].

Farmers who have experienced illness may also, understandably, face heightened challenges in maintaining a positive outlook towards PPE. Illness can lead to physical discomfort, diminished work capacity, and potentially increased economic strain due to healthcare expenses. Such circumstances may divert attention and resources away from investing in protective gear [34]. Additionally, farmers who have personally encountered health issues may possess a different perception of occupational risks, potentially underestimating the imperative nature of PPE [26].

Our study also finds an intriguing finding that needs to be interpreted cautiously, the knowledge on PPE use being associated with lower odds of positive attitudes towards its PPE use. This counterintuitive relationship prompts closer examination of potential underlying factors. Firstly, it is relatable that farmers with extensive knowledge of PPE may, paradoxically, become cognizant of its limitations or discomforts in practical application. This heightened awareness may lead to a more relaxed attitude towards its use. Additionally, the excess of knowledge may inadvertently lead to complacency, as farmers might overestimate their proficiency in implementing safety measures, potentially reducing the perceived necessity of consistent PPE adherence. Furthermore, it is crucial to consider that attitudes towards safety practices are influenced by a lot of factors. Socio-cultural norms, personal experiences, and contextual constraints play pivotal roles in shaping these attitudes [35,36].

The marital status of farmers, their years involved in farming, years of pesticide use, number of hours of pesticide exposed, methods of applying pesticides, and their attitude towards PPE use were all found to be associated with practice related to PPE use. The farmers who were married were more likely to wear PPE while working with pesticides which is consistent with the study conducted in Cameroon [30]. A plausible explanation for this might be that the married individuals have a heightened sense of responsibilities towards their family, having any form of illness not only affects the individuals but also adds financial burden to their family therefore they might adhere to adopt responsible behaviors like wearing PPE while handling pesticides. Likewise, their spouse and other family members may also encourage and pressurize them to adopt precautionary measures.

Further, the odds of using PPE were found to be substantially lower among the farmers who were involved in farming for more than 10 years. A similar finding was seen in a study conducted in India where the farmers who had mean 18 years old farming exposure were not following safety practices [29]. This could be because those farmers may have developed sense of confidence in their work and have less perceived risk in their work by underestimating the potential hazard of pesticide and have a false belief that they can manage the risk without the use of PPE.

However, unlike the years involved in farming, the farmers who were using pesticides for more than 10 years had higher odds of using PPE. The plausible reason behind this may be because the farmers who were using pesticides for a longer time might be more familiar with the potential health hazards or might have experienced side effects and may be more familiar about the safety measures to reduce the risk. Moreover, as farmers accumulate years of experience, they often become more attuned to industry recommendations and regulatory guidelines regarding safety practices [29]. Likewise, the farmers who were exposed to pesticides for 10 hours or more in their lifetime were more likely to use PPE. This might be because the farmers might perceive that an exposure to pesticides for a longer period can cause deleterious effects which has also been mentioned in theory of toxicology [29,31].

Additionally, the finding that positive attitude was associated with lower odds of PPE use among farmers presents an intriguing and potentially counterintuitive observation. A similar finding was also presented in a study conducted in Ethiopia [27]. This result may be elucidated by several factors outlined in existing literature. Firstly, individuals with an optimistic outlook towards their work environment may exhibit a heightened perception of safety and reduced perceived risk, potentially diminishing the perceived necessity for PPE. This phenomenon, known as the “optimism bias,” has been documented in various occupational contexts, where individuals tend to underestimate their personal susceptibility to negative events [32]. Moreover, positive attitude may inadvertently lead to overconfidence, as farmers may believe that their positive outlook alone is sufficient to mitigate potential risks, thereby diminishing their motivation to engage in precautionary measures like PPE use. Additionally, socio-cultural factors and peer influence may play a pivotal role in shaping attitudes towards safety practices; downplaying the necessity of PPE use [33,34]. This result underscores the importance of considering not only attitudes but also their potential impact on behavior. It emphasizes the need for targeted interventions that address the complex interplay between attitudes, risk perception, and actual safety practices to ensure optimal PPE utilization among farmers.

This study also has some limitations. Firstly, the interviews were conducted through telephone considering safety protocols during the COVID-19 pandemic; therefore, the responses are subjected to differ from face-to-face interviews as the communication might have been subject to various interference like lack of visual cues, body language and time constraint which might have influenced the data and the study. Further, the pandemic increased the use of masks among people and since this study is also related to the use of PPE, the situation may also have influenced the result of the current study.

## Conclusion

In conclusion, this study provides valuable insights into the factors influencing the adoption of PPE among farmers. The association between education and knowledge on PPE underscores the pivotal role of formalized education in promoting safety awareness and practices within the agricultural community. Additionally, the association between lower socioeconomic status and attitudes towards PPE highlights the need for targeted interventions to address economic barriers and ensure equitable access to protective gear. Furthermore, the finding that farmers who have experienced illness exhibit lower odds of positive attitudes towards PPE emphasizes the importance of considering health status as a determinant of safety behaviors. Moreover, the positive relationship between years of pesticide uses and increased PPE utilization highlights the evolution of safety consciousness over time, underlining the value of experiential learning in shaping responsible pesticide handling behaviors. This study has several implications for policy and practice. It underscores the need for tailored educational programs that consider the diverse educational backgrounds and experiences of farmers. Additionally, targeted interventions addressing economic constraints and health status are crucial in promoting consistent PPE use. Finally, efforts to foster a culture of safety within the agricultural sector should encompass not only knowledge dissemination but also the cultivation of positive safety attitudes.

## Data Availability

All relevant data are within the manuscript and its Supporting Information files.

## Acknowledgments

We would like to express our gratitude to Professor Shital Bhandary, School of Public Health, Patan Academy of Health Sciences for the support during data analysis. Additionally, we thank Kirtipur Municipality for allowing us to conduct the study.

## Notes

### Competing Interest Statement

The authors have declared no competing interest.

### Funding Statement

The author(s) received no specific funding for this work.

### Author Declarations

Ethical approval was obtained from the Institutional Review Committee (IRC) of Patan Academy of Health Sciences (PAHS)- PAHS IRC Reference PHP2107221558.

